# Assessment of Post SARS CoV 2 Fatigue among Physicians Working in COVID Designated Hospitals in Dhaka, Bangladesh

**DOI:** 10.1101/2021.02.08.21251352

**Authors:** A T M Hasibul Hasan, Muhammad Sougatul Islam, Nushrat Khan, Nazmul Hoque Munna, Wahidur Rahman Choton, Mostofa Kamal Arefin, Mohammad Abdullah Az Zubayer Khan, Mohaimen Mansur, Rashedul Hassan, Numera Siddiqui, Muhammad Shamsul Arefin, Nayema Afroze, Md. Salequl Islam

## Abstract

**Background:** Fatigue has been observed after a number of infectious disease outbreaks around the world. After the outbreak of SARS CoV-2 in Wuhan, China in 2019, the disease turned into a pandemic very rapidly. Mental health is a key issue associated with such outbreaks. To explore the fatigue level among physicians working in designated public and private hospitals in Bangladesh, we conducted a matched case-control study of post-SARS-CoV-2 fatigue.

**Method:** In this study 105 physicians who were diagnosed as COVID-19 infected, got treatment, and declared cured at least 6 weeks before the interview date, were recruited as cases and the same number of age and designation matched healthy physicians as control who are working in the same hospital. Case and control were selected in 1:1 ratio from each of the hospitals. The study population was selected by inclusion and exclusion criteria after taking informed written consent. Data collection was done by a semi-structured questionnaire. Diagnosis of COVID--19 infection was done by detection of SARS CoV-2 antigen by RT-PCR from reference laboratories in Bangladesh or by HRCT Chest.

**Result:** Around two-thirds of the physicians were male (67.6% versus 32.4%). Most of them aged less than forty years (80.5%). The cases had a greater number of comorbid conditions than those who were negative. The FSS score (mean) was much higher for cases (36.7 ± 5.3 versus 19.3 ± 3.8) than the control group with a statistically significant difference with no significant gender differentiation. Similarly, around 67.7% of the previously COVID positive physicians represented in the highest FSS score tertile compared to the respondents in the control group had a mean score of less than 3. The difference was also highly significant.

**Conclusion:** Physicians, who had a previous history of COVID-19 infection had a higher total and mean FSS score, signifying a more severe level of fatigue than the physicians who had never been COVID-19 positive while working in the same hospital irrespective of their age and sex.

## Introduction

COVID 19 has created an unprecedented public health emergency for almost a year since its emergence from Wuhan, China in December 2019 (Novel Coronavirus – China, 2021). With the alarming rate of infectivity, considerable rate of mortality, and privation of effective treatment as well as an effective vaccine to prevent transmission, has made the disease the biggest health and economic threat of the century (Fiani et al., 2020). Although initially emerged as a respiratory illness, COVID-19 was later established to cause more widespread symptomatology for its affinity to ACE-2 receptors (Carod Artal, 2020). It is nearly a year since the first-ever COVID-19 case was detected in Bangladesh on 8^th^ March 2020. Initially, the number of case detection was scarce. But with the sharp upsurge in case numbers, Bangladesh exceeded more than five hundred and seventy thousand cases by the end of January 2021 (WHO, 2021).

Presenting symptoms in our population were fever, cough, headache, myalgia, sore throat, malaise, and respiratory distress similar to all affected countries in the world (Ahmed et al., 2020. Experience from recent pandemics of severe acute respiratory syndrome (SARS) and the Middle East respiratory syndrome (MERS) virus points to potential long-term sequelae of COVID-19 that we might have to deal with in near future (Fraser, 2021). Several studies have already suggested that COVID-19 might not only leave a scar in the lung but also have a wide range of sequelae involving the cardiovascular and neuro-psychiatric system (Vindegaard and Benros, 2020), (Ojha et al., 2020).

In terms of mental health conditions, various research revealed a huge number of cases of depression, anxiety, fatigue, and post traumatic stress after the surge of SARS CoV-1 and MERS (Vindegaard and Benros, 2020). Lee AM et al observed that more than a third of the hospitalized SARS CoV-1 patients suffered from moderate to severe symptoms of depression and anxiety (Lee et al., 2007). Not only in the general population but post-pandemic/ post epidemic mental health conditions of healthcare workers and doctors have also been documented in various research. Moldofsky H et al have reported a syndrome of chronic fatigue, pain, weakness, depression, and sleep disturbance, which overlaps with the clinical and sleep features of fibromyalgia syndrome and chronic fatigue syndrome among health care workers after SARS CoV-1 in his case-control study (Moldofsky and Patcai, 2011).

Health care providers are also particularly vulnerable to emotional distress in the ongoing COVID-19 pandemic, given their risk of exposure to the virus, concern about infecting and caring for their loved ones, shortages of personal protective equipment (PPE), longer work hours, and involvement in emotionally and ethically fraught resource-allocation decisions (Pfefferbaum and North, 2020). A recent study on Chinese doctors and nurses reported most of the doctors and nurses working in COVID-19 designated departments have a very high prevalence of psychological distress, anxious symptoms, and depressive symptoms (Liu et al., 2020). Long working hours along with all these factors can lead to fatigue especially burnouts. As for Bangladesh, the prevalence of anxiety symptoms and depressive symptoms was 33.7% and 57.9%, respectively, and 59.7% mild to extremely severe levels of stress among the general population of Bangladesh during the COVID-19 pandemic (Banna et al., 2020). Until now, there is no evidence on the mental health status of the healthcare workers of Bangladesh during the pandemic; which can further affect their personal life as well as professional commitments if not provided sufficient attention and required mental health services.

Therefore, we conducted this case-control study to assess the level of fatigue after COVID-19 infection among the physicians working under a similar working environment of COVID-19 designated hospital in Bangladesh and generate evidence on this unexplored scenario.

## Methodology

This matched case-control study was conducted between April 2020 to September 2020 among physicians working in ten COVID designated hospitals (both public and private) in Dhaka, Bangladesh. Data were collected from Dhaka Medical College Hospital, Mugda Medical College Hospital, Kurmitola General Hospital, Kuwait Maitree Hospital, Mymensingh Medical College Hospital, Sayed Nazrul Islam Medical College Hospital, Green Life Medical College, Holy Family Red Crescent Medical College Hospital and Anwar Khan Modern Medical College Hospital from working physicians through a structured questionnaire.

### Diagnosis and confirmation of SARS CoV-2 infection

Diagnosis of SARS CoV-2 infection was based on clinical presentation along with evidence of the presence of viral antigen by RT-PCR test through the throat and nasal swab; irrespective of any evidence of COVID-associated pneumonia in HRCT Chest.

### Case Definition

Physicians working either in a public or private COVID-19 designated hospital, who had a history of COVID-19 infection as evidenced by clinical feature and RT-PCR for SARS CoV-2 antigen from nasal and throat swab with or without HRCT Chest; and was declared cured by negative test result by RT-PCR at least six weeks before the interview date was defined as a case for this study.

Physicians who were asymptomatic but tested positive for COVID-19 by RT-PCR was not included in this study. Furthermore, physicians who had typical HRCT changes for COVID-19 infection but had negative RT-PCR test results were also not included in this study.

### Control Definition

In this study controls were healthy physicians, selected in 1:1 ratio from each of the study centers matching for both age and professional designation with the cases; and who had a negative report on RT-PCR for SARS CoV-2 infection in nasal and throat swab. This is to mention, all physicians working in COVID-19 designated hospitals are subjected to routine RT-PCR tests for SARS CoV-2 before and after their rotational ward duties.

### Sample size

It was a matched case-control study. We matched participants based on age and their professional hierarchy (to correct confounding for exposure level as it can be widely varied due to assigned duties). Matching was done on a 1:1 basis. As COVID-19 is a newly immerged clinical condition and therefore inadequate data for prevalence or odds ratios to calculate the exact required sample size, we adopted the traditional arbitrary method. So with an expected proportion amongst controls of 0.05%, assumed odds ratio of 4, 95% confidence interval, and 80% power, the required minimum sample size was 196 in total and 98 in each group. We increased the sample size to 210 with 105 participants each in case and control groups.

### Study site and sampling

Data were collected from 6 government and 4 private hospitals in Dhaka city; all of which were dedicated for COVID-19 t. A convenience sampling method was adopted to collect data for this study.

### Assessment of Fatigue- FSS Questionnaire

Fatigue can be defined as a subjective experience and includes such symptoms as rapid inanition persisting lack of energy, exhaustion, physical and mental tiredness, and apathy (Chaudhuri and Behan, 2004). The level of fatigue was assessed using the Fatigue Severity Scale (FSS) which has prior validation through various studies (Valko et al., 2008); which was developed by Krupp in 1989 (Armutlu et al., 2007). The FSS is a 9-item questionnaire for the assessment of the severity of Fatigue (Table 1). In this self-administered questionnaire grading of each item ranges from 1-7, where 1 is designated for strong disagreement and 7 for a strong agreement. The lowest score is 9 and the highest can be 63. The mean score of all items is also considered according to reporting/study methodology (Armutlu et al., 2007).

**Table 1:** Fatigue Severity Scale

| Fatigue Severity Scale (English) |  |  |  |  |  |  |  |
| --- | --- | --- | --- | --- | --- | --- | --- |
|  | strongly disagree<br>strongly agree |  |  |  |  |  |  |
|  | 1 | 2 | 3 | 4 | 5 | 6 | 7 |
| 1. My motivation is lower when I am fatigued. | 0 | 0 | 0 | 0 | 0 | 0 | 0 |
| 2. Exercise brings on my fatigue. | 0 | 0 | 0 | 0 | 0 | 0 | 0 |
| 3. I am easily fatigued. | 0 | 0 | 0 | 0 | 0 | 0 | 0 |
| 4. Fatigue interferes with my physical functioning. | 0 | 0 | 0 | 0 | 0 | 0 | 0 |
| 5. Fatigue causes frequent problems for me. | 0 | 0 | 0 | 0 | 0 | 0 | 0 |
| 6. My fatigue prevents sustained physical functioning. | 0 | 0 | 0 | 0 | 0 | 0 | 0 |
| 7. Fatigue interferes with carrying out certain duties and responsibilities. | 0 | 0 | 0 | 0 | 0 | 0 | 0 |
| 8. Fatigue is among my three most disabling symptoms. | 0 | 0 | 0 | 0 | 0 | 0 | 0 |
| 9. Fatigue interferes with my work, family, or social life. | 0 | 0 | 0 | 0 | 0 | 0 | 0 |
| *\*Patients are instructed to choose a number from 1 to 7 that indicates their degree of agreement with each statement where 1 indicates strongly disagree and 7, strongly agree. [Krupp et al, Arch Neurol 1989]* |  |  |  |  |  |  |  |

### Data Collection and analysis

Data were collected through a structured questionnaire including questions related to basic demographic characteristics of the respondents, COVID-19 related status, and questions related to individual fatigue assessment. The data collection tool was pretested and modified before actual data collection. Data were collected by the researchers themselves. Data analysis was done by STATA version 16. Both descriptive and inferential statistical tests were done for data analysis and presentation. The mean FSS score and FSS score (total) were compared among cases and control to see any difference in the level of fatigue between the two groups.

### Ethical Issue

The study protocol was reviewed and approved by the Biosafety, Biosecurity and Ethical Committee of Jahangirnagar University, Bangladesh [BBEC, JU/M 2020/COVID-19/(11)1]. Written informed consent was taken from each participant after providing a detailed explanation about the objective, methodology, and purpose of the study. Participants were also informed about no physical risk involved with the study along with data integrity. Also, it was made clear that no physical, psychological, social, legal, or other risk was present in the study and participants could withdraw anytime from the study.

## Result

In this study, a total of 210 age and designation matched COVID-19 positive and COVID-19 negative physicians were interviewed in a 1:1 ratio. Around two-thirds of the physicians were male (67.6% versus 32.4% female). Most of them aged less than forty years (80.5%). Although 74.5% of them had some form of postgraduate degrees, most of them were junior physicians working as a medical officer or assistant registrar (82.9%) (Table 2). Most of the participants worked in different government hospitals with an overall mean of 8.1 (SD 5.4) years. About 77.1% of the previously COVID-19 positive physicians (cases) were aged below 40 years whereas 83.8% in COVID-19 negative group were of the same age. All the differences were statistically nonsignificant between case and controls. For the number of pre-existing comorbidities, the difference was statistically significant. In terms of FSS score, it was relatively much higher in cases than in the control group. The mean value of the FSS score was also much higher and statistically significant in the case group (36.7 Vs 19.3).

**Table 2:** Study participant description

| Variables | Case |  | Control |  | p-value |
| --- | --- | --- | --- | --- | --- |
|  | n | % | n | % |  |
| **Age (years)** |  |  |  |  |  |
| <30 | 17 | 50.0 | 17 | 50.0 |  |
| 31-40 | 67 | 50.0 | 67 | 50.0 |  |
| 41-50 | 20 | 50.0 | 20 | 50.0 |  |
| 51-60 | 1 | 50.0 | 1 | 50.0 |  |
| **Sex** |  |  |  |  | 0.140 |
| Male | 66 | 46.5 | 39 | 57.4 |  |
| Female | 76 | 53.5 | 29 | 42.7 |  |
| **Postgraduate degree** |  |  |  |  | 0.004 |
| None | 16 | 29.6 | 38 | 70.4 |  |
| Diploma/MCPS/MPhil | 23 | 51.1 | 22 | 48.9 |  |
| MD/MS | 36 | 61.0 | 23 | 39.0 |  |
| FCPS | 30 | 57.7 | 22 | 42.3 |  |
| **Current designation** |  |  |  |  |  |
| Medical Officer/ Assistant Registrar/ Lecturer/Registrar | 87 | 50.0 | 87 | 50.0 |  |
| Junior Consultant/ Assistant Professor | 15 | 50.0 | 15 | 50.0 |  |
| Senior Consultant/Associate Professor | 2 | 50.0 | 2 | 50.0 |  |
| Professor | 1 | 50.0 | 1 | 50.0 |  |
| **Workplace** |  |  |  |  | 0.000 |
| Public Hospital/ Institute/ Medical College/University | 51 | 76.1 | 16 | 23.9 |  |
| Private Hospital/ Institute/ Medical College/ Clinic | 54 | 37.8 | 89 | 62.4 |  |
| **Comorbidity** |  |  |  |  | 0.002 |
| 0 | 48 | 45.7 | 50 | 47.6 |  |
| 1 | 27 | 25.7 | 14 | 13.3 |  |
| 2-5 | 26 | 24.8 | 21 | 20.0 |  |
| >6 | 4 | 3.8 | 20 | 19.1 |  |
| **FSS Score** |  |  |  |  | 0.000 |
| 1(lowest) | 1 | 1.4 | 72 | 98.6 |  |
| 2 | 38 | 53.5 | 33 | 46.5 |  |
| 3(highest) | 66 | 100.0 | 0 | 0.0 |  |
|  | Mean | SD | Mean | SD |  |
| **FSS Score** | 36.7 | 5.3 | 19.3 | 3.8 | 0.000 |
| **Years of the clinical career** | 8.4 | 5.7 | 7.9 | 5.2 | 0.5104 |

\*Significance after Bonferroni Correction

In table 3, gender differentials for various variables in COVID-19 positive participants are shown. Most of the participants were confirmed as COVID-19 positive cases by the RT-PCR test. Only a third of the participants required hospitalization with a male majority. Only 6 of the total cases required ICU support. In terms of self-perceived severity, more than 90% of cases were mild to moderate. Pre-existing comorbidities were higher in men, but none of these characteristics were statistically significant. Relative FSS score was higher among the male cases, although the mean score was almost similar in male and female, but statistically non-significant.

**Table 3:**
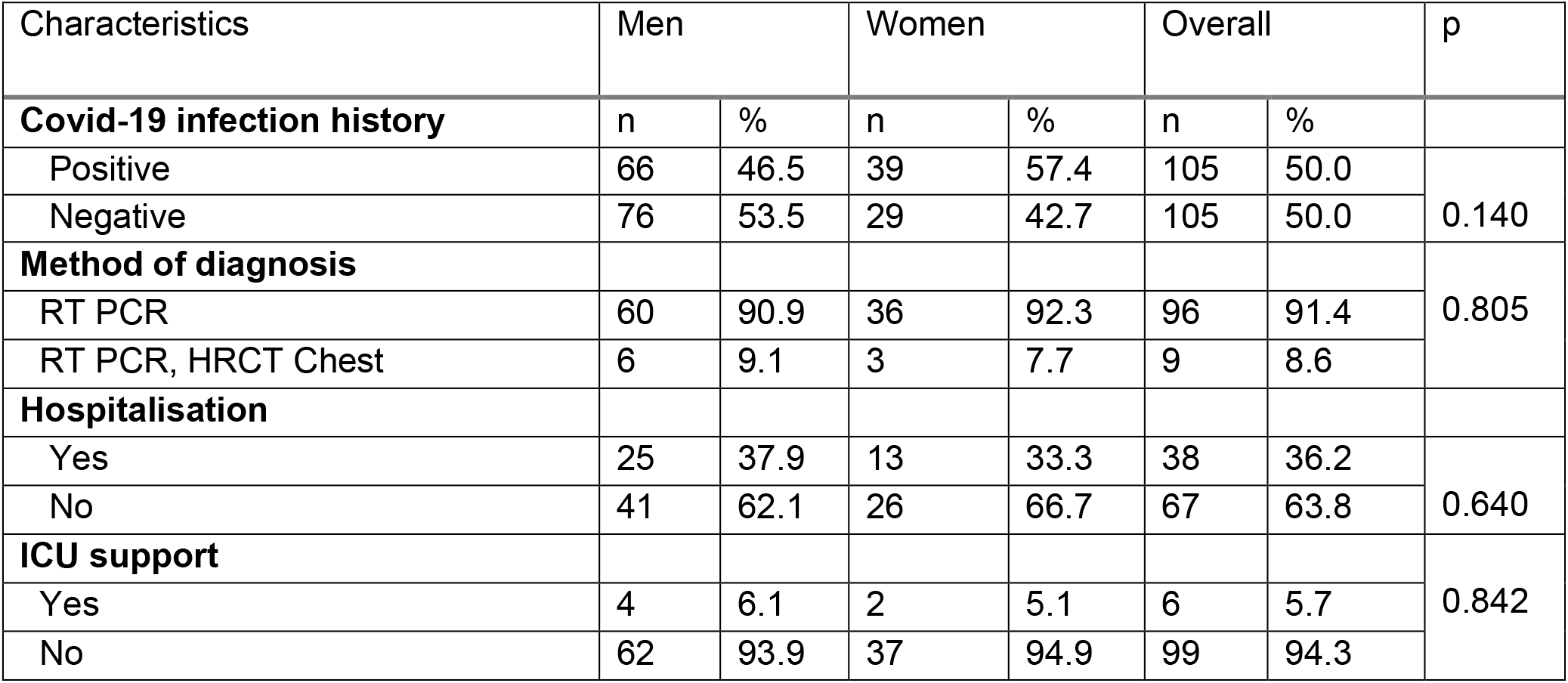
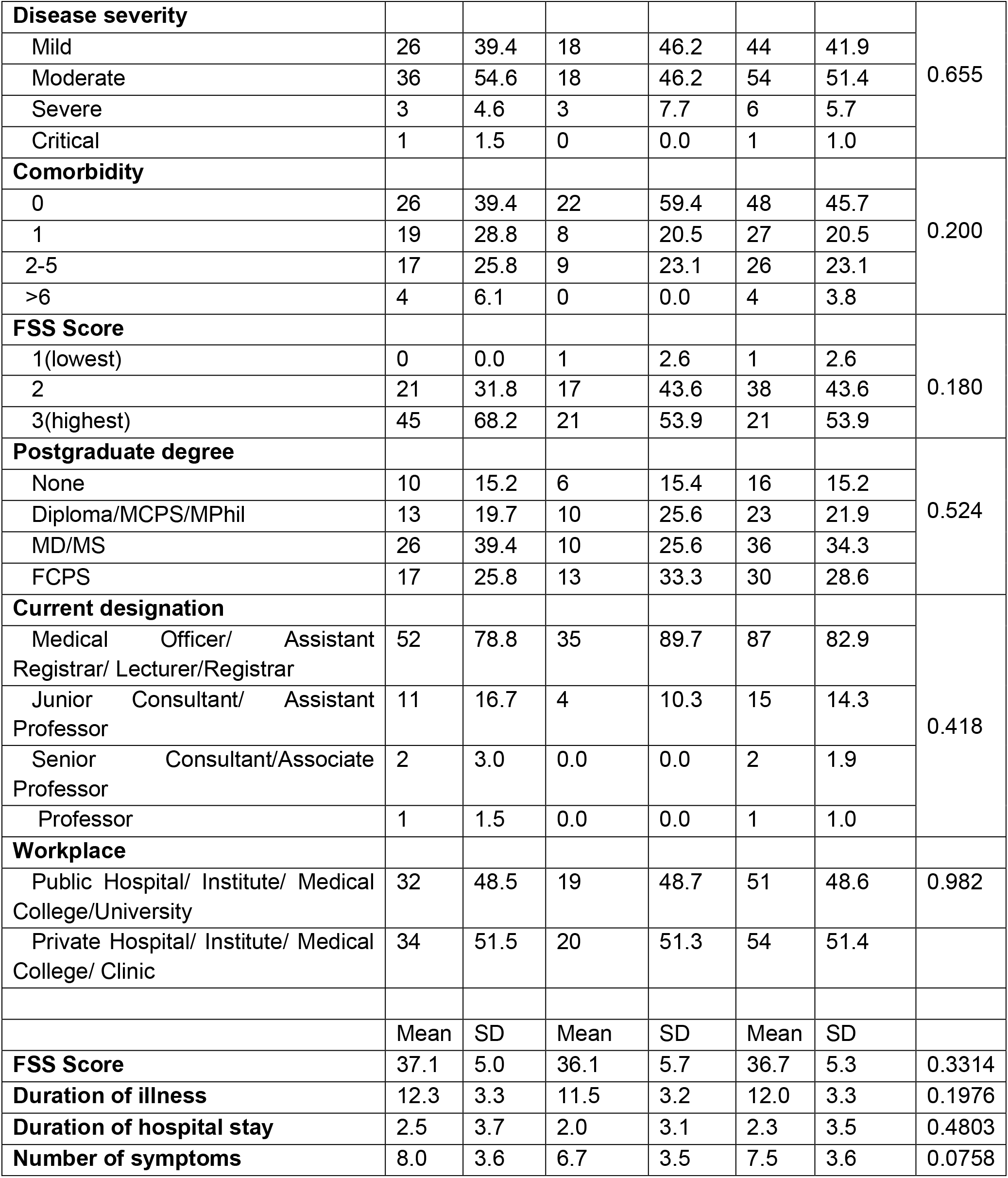
COVID-19 related characteristics of the study participants

**Table 4:** Multivariate analysis of predictor variables in cases compared with controls

| Characteristics | Model 1 | Model 2 | Model 3 |
| --- | --- | --- | --- |
| **Sex** |  |  |  |
| Male | 1 |  | 1 |
| Female | 1.5 (0.8-2.8) |  | 3.8 (0.6-23.3) |
| **Postgraduate degree** |  |  |  |
| None | 1 | 1 | 1 |
| Diploma/MCPS/MPhil | 2.5 (1.1-5.7) * | 2.4 (1.1-5.6) * | 0.4 (0.0-3.8) |
| MD/MS | 3.7 (1.7-8.1) * | 3.8 (1.7-8.3) * | 2.3 (0.2-22.0) |
| FCPS | 3.2 (1.5-7.2) | 3.2 (1.4-7.1) * | 0.1 (0.0-1.8) |
| **Workplace** |  |  |  |
| Private Hospital/ Institute/<br>Medical College/ Clinic | 1 | 1 | 1 |
| Public Hospital/ Institute/<br>Medical College/University | 5.3 (2.7-10.1) * | 5.2 (2.7-10.1) * | 6.7 (0.7-68.0) |
| **Comorbidity** |  |  |  |
| No | 1 | 1 | 1 |
| Yes | 1.1 (0.6-1.9) | 1.1 (0.6-2.0) | 0.7 (0.1-5.1) |
| **FSS Score** | 1.8 (1.5-2.1) * | 1.8 (1.5-2.1) * | 2.0 (1.5-2.7) * |

Model 1: Unadjusted; Model 2: Adjusted sex; Model 3: Fully adjusted

**Figure 1:**
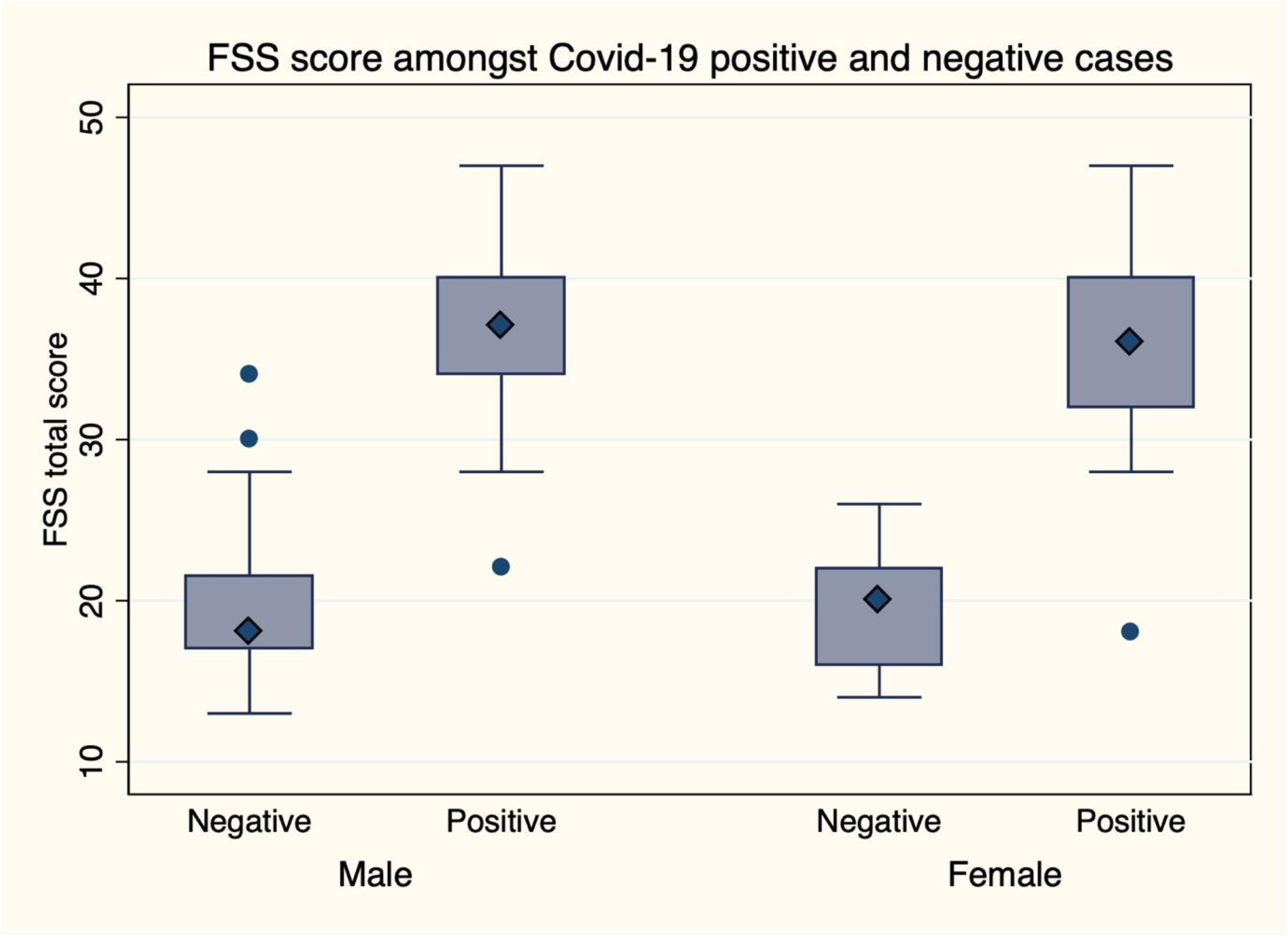
FSS Score among case and controls

Next, in the multivariate logistic regression 3 models were implemented. There was no significant association between demographic and professional characteristics between cases and controls. But in terms of FSS score, the odds of developing post-covid-19 fatigue was two times more in cases than in controls, which was significant in unadjusted, sex-adjusted, and fully adjusted models.

**Figure 2:**
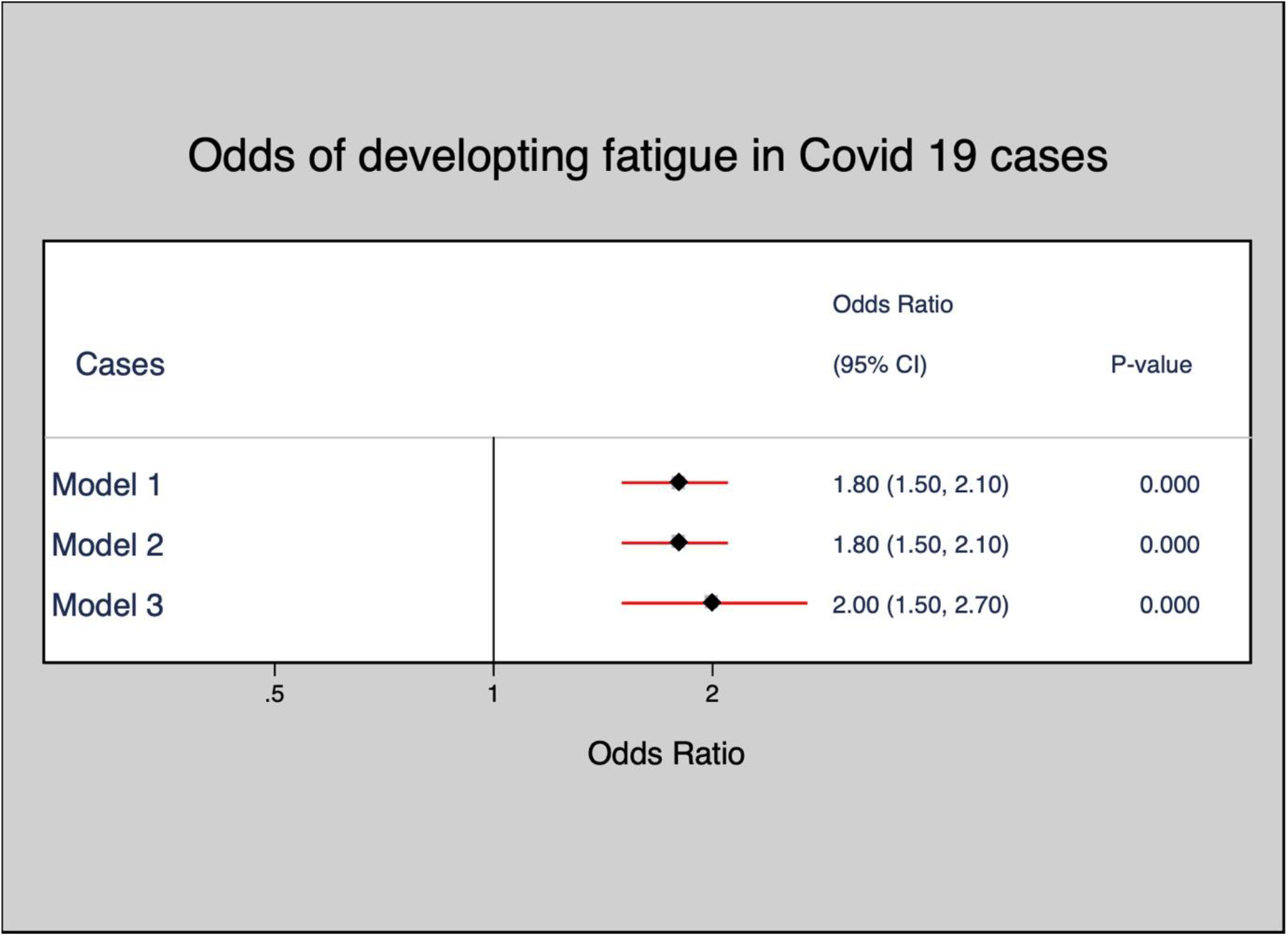
Odds of developing post-covid-19 fatigue in the cases

## Discussion

In this case-control study of post-COVID-19 fatigue among physicians working in COVID-19 designated hospitals in Bangladesh, we noted a higher FSS score (both relative and mean) in previously COVID-19 positive physicians as compared to never positive physicians. However, we did not find evidence that there was any difference between cases and controls in terms of their sex, postgraduation degrees, number of years of service, or pre-existing comorbid conditions.. In fact, the difference in scores from cases and indicated an increasing level of fatigue in people who have been positive with the virus infection and now back to work station.

Our initial literature review of major medical databases revealed a considerably low number of studies focusing on the mental health status of health care workers engaged directly in the diagnosis and treatment of patients infected with SARS CoV-2. However, we could find only one study that emphasized the mental health status of health care workers who previously contracted the COVID-19. Junhua et al, reported psychological factors and sleep status of medical staff contracting the COVID-19. In their case-control study, they assessed seventy medical staff for mental health status who had COVID-19. They used Symptom Checklist-90 (SCL-90), Patient Health Questionnaire (PHQ-15), Self-rating Anxiety / Depression Scale (SAS/ SDS), and Post-Traumatic Stress Disorder self-assessment scale (PTSD Checklist-Civilian Version, PCL-C) and Pittsburgh Sleep Quality Index (PSQI) for assessment of mental health. Those who were infected previously with COVID-19 had significantly higher somatization, depression, anxiety, phobia, and sleep disturbances than the control group (MEI, Zhang, Gong, and Lijuan, 2020). This resonates with the fact of this understudied condition until now. Psychiatric comorbidities and persistent mental health problems had been reported in both short-term and long-term follow-up studies after the pandemic of SARS CoV-1 in 2003 (Tansey, 2007; Lee et al., 2007; Lam, 2009). Chronic Fatigue Syndrome (CFS) had been reported in around 40.3% of survivors of SARS CoV-1 (Lam, 2009). The strength of our study is, it is the first study in South Asia to explore this area of interest and with the finding of significantly high FSS score in cases, sets the base for further exploration.

The long-term effect of SARS CoV-2 is yet to be established. Undue tiredness, irritability, sleep disturbance, fatigue, and other different somatic symptoms had been reported throughout the world after previous outbreaks of various epidemics/pandemics. These are often termed as post-infectious fatigue syndrome (PIFS) or post-viral fatigue syndrome (PVFS) (Naess et al., 2012; Hickie et al., 2006; Hickie et al., 2009; Behan and Bell, 1985).

The global prevalence of PIFS ranges from 0.2% to 0.42%, which has a substantial economic impact with the loss of productivity and loss of employment at the individual level (Nacul et al., 2011; Jason, Taylor and Kennedy, 2000; Lin et al., 2011). The economic costs generated by chronic fatigue are high and mostly borne by patients and their families. Loss of income leads to a reduced standard of living, increase the health care cost and ultimately increase the expenditure budget on the respective Government (Collin et al., 2011; Jason et al., 2008; Lloyd and Pender, 1994; Lin et al., 2011; Sabes-Figuera et al., 2010).

Therefore, this study was conducted on a relatively smaller scale to understand the impact of COVID 19 infection among physicians in Bangladesh working in COVID-19 designated hospitals. Inevitably, the majority of the physicians (80.5%) in this study were below forty years of age who are the main driving force of the health care system of Bangladesh. Around two-thirds of them were male physicians. Reasonably enough the cases had a greater number of comorbidities. In light of our observation, KT Bajgain et al, in his review of 22 studies involving 22,753 COVID cases around the world reported a higher prevalence of one or more comorbidities that includes CVD (8.9%), HTN (27.4%), Diabetes (17.4%), COPD (7.5%), Cancer (3.5%), CKD (2.6%), and other (15.5%) (Bajgain, Badal, Bajgain and Santana, 2021).

The 9 item FSS questionnaire was used for assessment of fatigue among the case and control groups. The cases had a significantly higher (p-value 0.000) mean FSS score (36.7 ± 5.3 versus 19.3 ± 3.8) and a higher relative FSS score in higher tertiles. This denotes that, although the physicians working under a similar stressful condition in COVID-19 designated hospitals irrespective of their age and designation; those with a previous history of contracting COVID-19 infection had significantly more severe fatigue than their colleagues who had never been positive by RT-PCR for SARS CoV-2 in nasal and throat swab test.

The mental health status of health care workers had been a great challenge since the outbreak of previous global pandemics; many times they had a long-term impact on the productivity and health status of the frontliners (Chong et al., 2004; Wu et al., 2009). For COVID-19, already studies have documented front line health workers directly involved with COVID patients had a higher risk of depression, anxiety, insomnia, and distress (Lai et al., 2020), our findings with post-infection fatigue is similar. To reduce respondent bias and influence of stress level at work we had compared the fatigue status of physicians working in different hospitals matching their designation. Also, the study was conducted at the peak stage of the pandemic, therefore recall bias on the part of participants should be minimal.

Our study is not free from limitations. First of all, there may be selection bias on relatively young physicians since the proportion of young physicians working in COVID-19 designated hospitals are much higher than the seniors. We tried to correct this by age and designation matching. Moreover, senior physicians aged more than 50 are not directly involved with the management of COVID-19 patients. The senior consultant physicians were supervising the COVID-19 management mostly online. So, we cannot infer the fatigue and comorbidity status of elder/senior physicians. Various other confounding factors might have been unexplored, e.g. environmental exposure, economic status, family history, etc. Finally, although case-control study designs are efficient for rare diseases, a cohort study could yield multiple outcomes and stronger associations; which was not possible due to time and financial constraints.

## Conclusion

Physicians who became COVID positive while working at COVID 19 designated hospital had a more severe level of fatigue at least six weeks post-infection than their COVID negative colleagues working in the same hospital in Bangladesh. This is evident by a significantly higher relative and means FSS score. Although it is still too early to comment whether there would be a similar or higher rate of PIFS or CFS among SARS CoV-2 survivors than those of SARS CoV-1; this study puts insight into the future mental health burden of these patients and recommends further large-scale research nationally.

## Data Availability

Data will be available through email

## Author contribution

Dr. ATM HH was involved in planning the study, designing study methodology, consultation and data collection, and manuscript writing for this study. NK was involved in data analysis, data interpretation, and manuscript writing. MSI was involved in data collection and editing the manuscript. All other authors were involved in data collection. All the authors have read and approved the final version of the manuscript.

## Acknowledgments

We acknowledge the contribution of our supporting staff at each of the public and private hospitals for helping us with the necessary data for the study.

